# Neural signatures of real-world turning during naturalistic locomotion in Parkinson’s Disease

**DOI:** 10.64898/2026.05.03.26352320

**Authors:** Rithvik Ramesh, Jannine Balakid, Jacob H. Marks, Kenneth H. Louie, Seongmi Song, Poojan Shukla, Hamid Fekri Azgomi, Doris D. Wang

## Abstract

Turning is a complex motor behavior that frequently triggers freezing of gait and falls in Parkinson’s disease (PD), yet its neural dynamics in naturalistic settings remain unknown. Using chronic at-home intracranial recordings in four subjects with PD, we show that turning is marked by premotor cortical beta desynchronization driven by reduced burst rate. These findings identify a robust signature of ecological turning and implicate beta dynamics in adaptive motor transitions.

## MAIN

Humans execute approximately 700 turns per day, each requiring coordinated integration of sensory input, postural control, and asymmetric motor output to reorient the body.^1,2^ ^3–6^ In Parkinson’s disease (PD), this common behavior becomes a major source of disability.^7–9^ Turning is often fragmented into a rigid ‘en bloc’ maneuver and frequently triggers for freezing of gait (FoG), markedly increasing fall risk.^10–12^ Current therapies have inconsistent effects on these axial symptoms. While dopamine replacement therapy may improve linear gait parameters such as speed, it can impair postural stability.^13–15^ Likewise, the effects of deep brain stimulation (DBS) on balance are highly variable.^16^ As a result, turning represents a vulnerable motor transition and a critical, yet under-addressed, therapeutic target.

Despite its importance, the neurophysiology of turning remains poorly understood. Conventional modalities are limited in studying this natural motor behavior; functional MRI requires immobility, while mobile EEG is constrained by motion artifacts and limited spatial resolution. ^17–20^ Moreover, laboratory-based paradigms lack ecological validity, capturing only brief, structured movements that fail to reflect the spontaneous, goal-directed nature of real-world locomotion.^2^ Tasks such as treadmill walking do not reproduce the dynamic postural adjustments or non-linear trajectories of everyday navigation, where turns are self-initiated and context-dependent.

To address this gap, we combined chronic, at-home intracranial recordings with wearable kinematics to define the neural dynamics underlying a key motor transition, turning, during naturalistic behavior in PD. Four subjects with PD (2M, 2F; Age: 62-68 years) with gait dysfunction (Unified Parkinson’s Disease Rating Scale Postural Instability and Gait Difficulty subscore; UPDRS PIGD Range: 2-10) were implanted with a bidirectional neurostimulator (Summit RC+S, Medtronic), with cortical and pallidal electrodes placed unilaterally (N=2) or bilaterally (N=2) (**Fig. 1a–c**).

**Figure 1.**
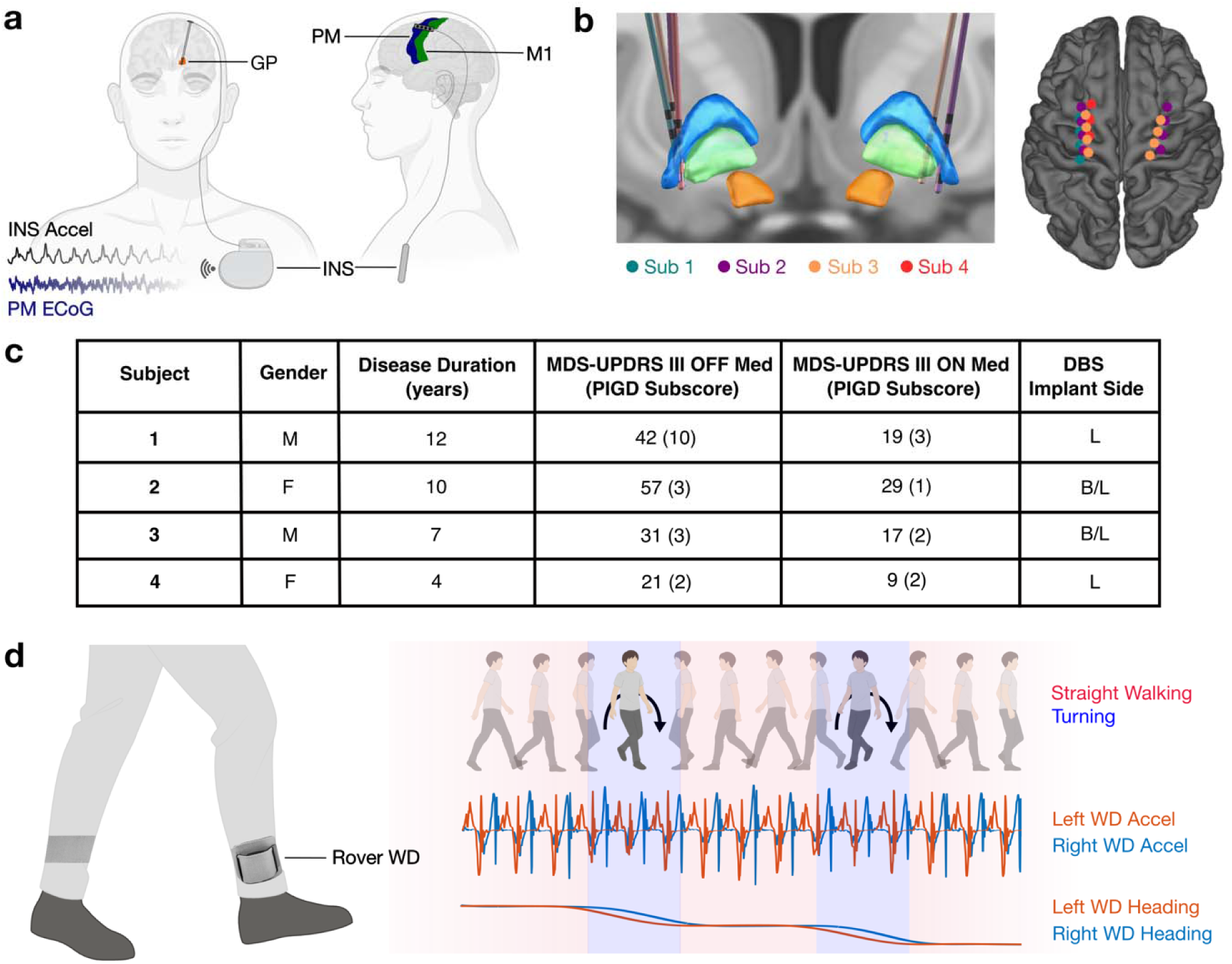
Overview of device and subject characteristics. **a**, Schematic of depth electrode implanted into GP and surface electrodes overlying M1 and PM, connected to Summit RC+S INS implanted superficially overlying ipsilateral pectoralis muscle. Sample acceleration and cortical ECoG signals wirelessly streamed from INS are shown. **b**, Depth and surface electrode reconstructions are shown for all subjects. **c**, Table showing subject age, gender, disease duration and severity, and DBS implant side. **d**, Schematic of Rover WDs, worn in fabric straps around the bilateral ankles (left). Sample subject activity synchronized with WD acceleration and heading signals is shown (right). Illustrations in subfigures **a** and **d** were partially created in BioRender. Ramesh, R (2026). GP, globus pallidus; M1, primary motor cortex; PM, premotor cortex; INS, implantable neurostimulator; LFP, local field potential; MDS-UPDRS, Movement Disorders Society Unified Parkinson’s Disease Rating Scale; PIGD, postural instability and gait difficulty; DBS, deep brain stimulation; L, left; R, right; B/L, bilateral; WD, wearable device.

To capture behavior in the home environment, participants wore lightweight ankle-mounted sensor devices (Rover, Sensoplex) integrating triaxial accelerometer, gyroscope, and magnetometer data. Foot orientation was derived from quaternion signals and converted to Euler angles to obtain the absolute heading, or yaw, for each foot. Subject-specific thresholds were applied to angular velocities calculated from each leg to enable labeling of turning and straight-walking epochs (**Fig. 2a,b**; **Methods**).

**Figure 2.**
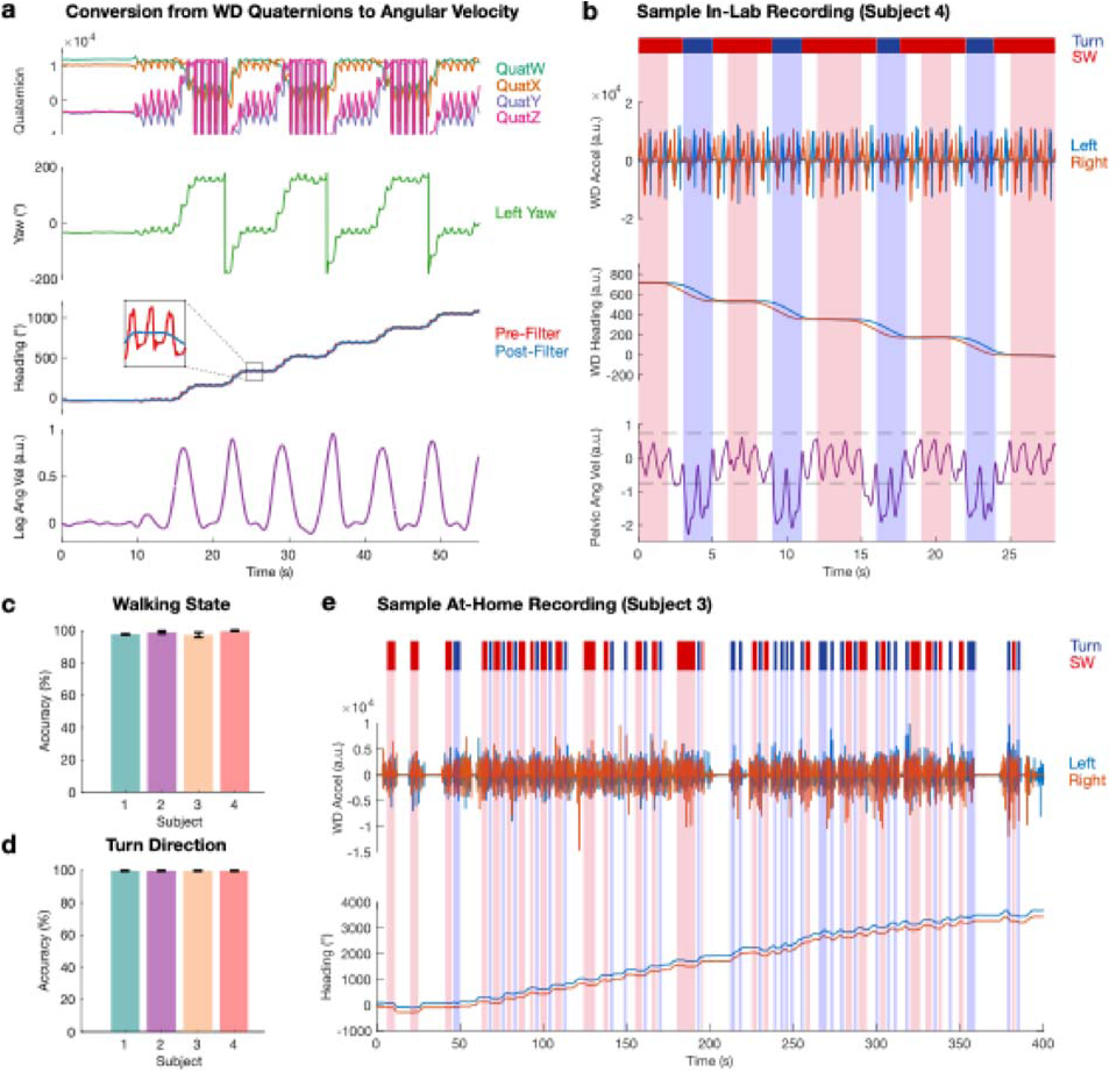
Wearable devices enable accurate labeling of turning and straight-walking. **a,** Sample raw quaternion signals are shown from a left wearable device (WD). Yaw signals were obtained by converting quaternions to Euler angles, which were then unwrapped and filtered to calculate a usable heading signal for each leg. Angular velocity for each leg was calculated as the first derivative of these heading signals. **b**, Sample in-lab WD labeling validation session is shown (Subject 4). Bar at the top indicates the true activity of the subject (blue, turning; red, straight-walking). Acceleration and heading signals are shown from left and right WDs, along with angular velocity from pelvic inertial measurement unit (IMU). Light blue and red overlays indicate WD-based turning and straight-walking labels respectively, showing excellent concordance with true walking states. **c**, Accuracies of WD-based walking state labeling are shown for all subjects. Bars show mean accuracy and error bars indicate standard deviation (SD). **d**, Accuracies of WD-based turn direction (i.e., left vs right) labeling are shown for all subjects. Bars show mean accuracy and error bars indicate SD. **e**, Sample at-home recording is shown. Acceleration and heading signals for bilateral WDs are shown. Light blue and red overlays indicate WD-based turning and straight-walking labels respectively. Quat, quaternion; WD, wearable device; SW, straight-walking; IMU, inertial measurement unit.

To validate wearable device (WD) based classification of gait states, we compared heading-derived labels of turning or straight-walking to ground-truth states defined by pelvic angular velocity during in-laboratory trials (see **Methods** for further details. A sample trial is presented in **Fig. 2b**. WD labels were highly accurate for all subjects, demonstrating accuracies ranging from 97.4% to 100.0% **(Fig. 2c; Supplementary Fig. S1)**. Sensitivity varied from 95.8-100.0% and specificity was 96.5-100.0%. Turn direction (i.e., left vs right) was accurately identified in all cases **(Fig. 2d**; **Supplementary Fig. S1)**. These results establish the reliability of wearable device kinematics for distinguishing walking states.

We next recorded multi-site neural and kinematic data during naturalistic behavior at home across a range of 5 to 12 days per subject **(Supplementary Table S1)**. Subjects remained on their clinically optimized stimulation settings and standard pharmacological regimens and did not receive any specific instructions guiding their movement in order to capture naturalistic behavior (example recording shown in **Fig. 2e**). Across six hemispheres, a total of 59.0 hours of synchronized neural-kinematic data were recorded, comprising 5,599 1-second turning epochs and 19,960 straight-walking epochs **(Supplementary Table S1)**. These data demonstrate the feasibility of capturing extended, unconstrained locomotor behavior using chronic, multi-site neural interfaces in the home environment.

To identify neural features distinguishing gait states, we first compared average spectral power across canonical frequency bands. Notably, premotor (PM) beta (13-30 Hz) power was consistently lower during turning compared to straight-walking across all six hemispheres (p<0.001) **(Figure 3a,b**, **Supplementary Figure S2**, and **Supplementary Table S2)**. Similar reductions in primary motor cortex (M1) alpha (8-13 Hz) and beta power were observed in most (4 of 6) hemispheres, though less consistently (p<0.01) **(Figure 3c)**.

**Figure 3.**
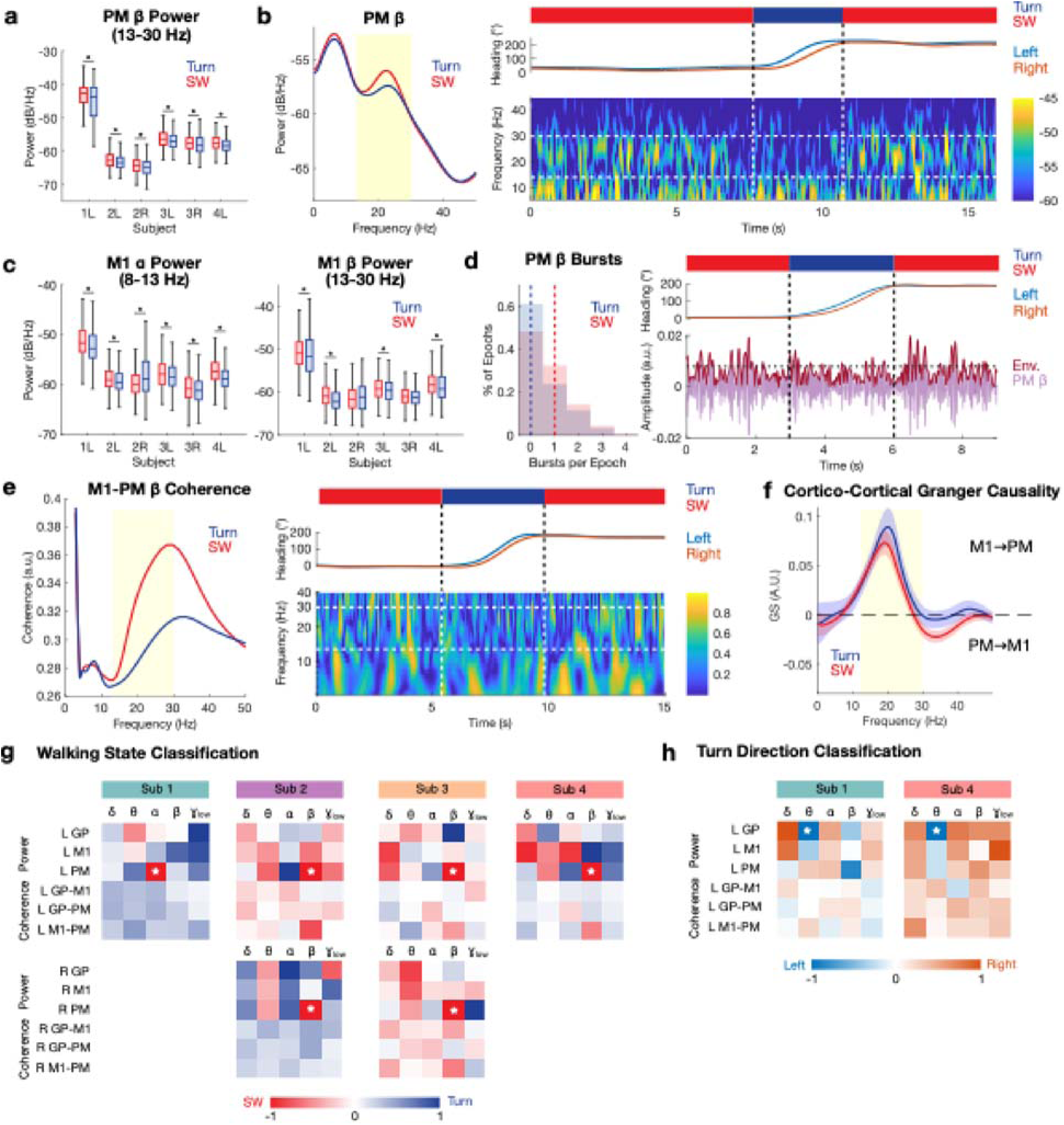
Cortical dynamics distinguish at-home turning and straight-walking. **a**, PM beta (13-30 Hz) power compared between all turning and straight-walking epochs within each hemisphere (two-sided Wilcoxon rank-sum tests with Benjamini-Hochberg correction). Box plots represent median, first quartile, and third quartile. Whiskers extend to non-outlier maximum and minimum (defined as falling within 1.5 x IQR). Asterisks indicate significant comparisons (p<0.05). **b**, Left, sample PSD is shown comparing average PM power between all turning and straight-walking epochs for a single hemisphere (Subject 4 Left). The shaded yellow region highlights the beta range, where a distinct difference is observed between walking states. Right, a sample of at-home turning and straight-walking behavior is shown for the same subject. The activity bar at the top indicates the behavioral state (blue, turning; red, straight-walking). The middle plot shows synchronized heading signals from the bilateral WDs. A spectrogram at the bottom shows temporal evolution of power across frequencies; vertical dashed lines delineate the turn period and horizontal dashed lines highlight the beta frequency band, showing decreased PM beta power during turning. **c,** Left, M1 alpha (8-13 Hz) and right, beta power compared between turning and straight-walking epochs within each hemisphere. **d,** Left, distribution of the number of PM beta bursts per epoch is shown for one hemisphere (Subject 3 Left). Dashed vertical lines indicate the median burst count for each walking state, showing decreased burst frequency during turning compared to straight-walking. On the right, a sample at-home segment is shown. Activity bar (top) and WD heading signals (middle) are shown, synchronized with a beta-filtered PM signal and its envelope (bottom). The horizontal dotted line represents the threshold used for burst detection. **e**, Left, average M1-PM coherence is shown for turning and straight-walking epochs in one Hemisphere (Subject 2 Left). Right, a sample at-home segment showing reduced M1-PM beta range coherence during turning. **f**, Average time-reversed Granger causality (TRGC) is compared between turning and straight-walking epochs. Positive scores indicate higher M1 to PM information flow, whereas negative scores indicate the opposite. **g,** Heatmaps displaying normalized logistic regression model coefficients (turning vs straight-walking) for each hemisphere. White asterisks highlight the importance of lower PM beta power during turning in most hemispheres and PM alpha power in the remaining hemisphere (Subject 1 Left). **h**, Heatmaps displaying normalized logistic regression model coefficients (left vs right turns) for the two hemispheres where above-chance decoding of turn direction was achieved. WD, wearable device; PM, premotor cortex; SW, straight-walking; M1, primary motor cortex; Env, envelope; TRGC, time-reversed granger causality.

To investigate the mechanism underlying state-dependent PM beta power suppression, we analyzed beta burst dynamics. Turning was associated with a reduced number of beta bursts per epoch in all but one hemisphere (Subject 2 left), without systematic changes in burst amplitudes and durations **(Fig. 3d** and **Supplementary Table S3)**. These findings indicate that beta modulation during turning is driven primarily by changes in burst rate rather than burst morphology.

We next examined inter-regional communication during turning and straight-walking periods. Differences in cortical-pallidal and cortical-cortical coherence between gait states were limited, although reduced M1-PM beta coherence was observed in a subset of hemispheres (Subject 2 left and Subject 4 left), **(Supplementary Figure S3** and **Supplementary Table S4)**. Directed connectivity analyses revealed consistent M1→PM information flow across both turning and straight-walking states, predominantly within the beta and low gamma (30-50 Hz) bands **(Supplementary Figure S4)**. Similar patterns were observed between in-laboratory and home recordings.

Having identified several differences in power and coherence during turning, we sought to isolate the most important neural features distinguishing turning from straight-walking. Individual logistic regression models were developed for each hemisphere, utilizing 30 distinct canonical power and coherence band features to differentiate gait states. Above-chance performance was achieved in all hemispheres (AUC: 0.56-0.77; p≤0.048; **Supplementary Table S5**), with reduced PM beta power emerging as the dominant feature in five of six hemispheres (**Fig. 3g; Supplementary Table S5**). In the remaining case (Subject 1 left), reduced PM alpha power during turning was the most important feature. Turn direction decoding was successful in two hemispheres (Subject 1 left and Subject 4 left), where pallidal theta (4-8 Hz) power was the most informative feature (**Fig. 3h** and **Supplementary Table S6**). Together, these results identify cortical beta desynchronization as the most robust neural signature of the transition from straight-walking to turning, with directional aspects potentially encoded by pallidal low-frequency dynamics.

Using chronic, at-home intracranial recordings, we identify premotor cortical beta desynchronization as a consistent neural signature of real-world turning in PD. This effect was observed across subjects and driven primarily by a reduction in beta burst rate, rather than changes in burst amplitude or duration, suggesting a specific modulation of transient inhibitory dynamics during behavioral transitions. These findings extend prior work linking beta suppression to movement by demonstrating its role in a complex, naturalistic motor transition that is highly relevant to disease-related disability. The consistency of this signal across unconstrained, real-world behavior highlights premotor cortex as a key node in enabling adaptive locomotor control. More broadly, our results suggest that cortical beta dynamics may provide a biologically grounded control signal for adaptive neuromodulation targeting axial symptoms in Parkinson’s disease.

Beta desynchronization has long been associated with motor control, with decreases in cortical beta power observed during movement and followed by post-movement rebound.^21–23^ This transient suppression is thought to reflect a release from the steady-state ‘status quo’, enabling flexible integration of sensorimotor inputs and adaptation of motor output .^24–27^ Consistent with this framework, prior studies in healthy individuals have shown reduced cortical beta activity during unconstrained compared to stabilized walking, supporting a role in balance and adaptability.^28,29^ In PD, this beta activity is characteristically hypersynchronized, leading to impaired movement flexibility. Despite this underlying pathological state, however, our findings demonstrate that task-related beta attenuation remains at least partially preserved during naturalistic turning and may represent a fundamental requirement for motor adaptation. Notably, we show that this suppression is mediated by reduced beta burst rate rather than changes in burst morphology, suggesting that modulation of discrete inhibitory events may increase temporal windows for sensorimotor processing and facilitate flexible motor control. ^30,31^

The limited ability to successfully decode turn direction may reflect variability in electrode placement or the transient nature of directional encoding, which may not be fully captured by epoch averaging. The identification of pallidal theta activity as a feature associated with turn direction is consistent with prior work implicating the globus pallidus in movement initiation and scaling, particularly for asymmetric muscle activation and leg trajectories during turns.^32–35^ These findings suggest that while cortical beta dynamics robustly encode state transitions, subcortical low-frequency activity may contribute to finer aspects of movement specification.

These results have important implications for neuromodulation. Emerging frameworks emphasize the need to target symptom-specific neural networks to optimize therapeutic outcomes.^36^ Given the anatomical and functional segregation of gait-related networks, understanding the neurophysiology of axial symptoms is essential for improving therapy, and identifying biomarkers of axial motor states is critical for advancing adaptive deep brain stimulation.^37,38^ Our findings suggest that cortical signals, particularly premotor beta dynamics, may serve as reliable indicators of behaviorally relevant transitions and could be leveraged to guide state-dependent stimulation. This supports a paradigm in which cortical recordings complement or extend traditional subcortical biomarkers, particularly for complex behaviors such as turning, gait initiation, and FoG.^39–41^

More broadly, this work highlights the importance of ecological validity in human neuroscience. Neural activity recorded during constrained laboratory tasks may not fully capture the dynamics of real-world behavior, which is inherently spontaneous, context-dependent, and cognitively demanding. By leveraging chronic at-home recordings, we establish a framework for studying brain activity during natural behavior, supporting both mechanistic insight and potential translational application. Such approaches may help bridge the gap between laboratory discovery and therapeutic implementation.

## METHODS

### Subject identification and DBS implantation

Four people with idiopathic Parkinson’s disease (PD) were recruited from those being evaluated for deep brain stimulation (DBS) implantation at the University of California, San Francisco. Subjects were implanted unilaterally in the left hemisphere (N=2) or bilaterally (N=2) with a novel bidirectional neurostimulator (Summit RC+S, Medtronic, Inc.; **Fig. 1a**). Depth leads were placed into the globus pallidus (GP) (Model 3387, Medtronic, Inc.; contact length: 1.5 mm; intercontact distance: 1.5 mm) and subdural quadripolar paddles were positioned over the sensorimotor cortices (Model 0913025, Medtronic, Inc.; contact diameter: 4 mm; intercontact distance: 10 mm) inserted through the same skull opening. Further details of the implantation surgery are available in prior papers.^42^ Electrode localization was performed using established image processing pipelines (Depth: Lead-DBS; Cortical: LeGUI; **Fig. 2b**).^43,44^ Briefly, postoperative computed tomography (CT) scans were coregistered to preoperative T1-weighted 3T magnetic resonance imaging (MRI) scans using a rigid linear affine transformation. Coregistration accuracy was verified manually and refined when necessary using an additional brain shift correction routine to align subcortical anatomy. Subcortical electrode artifacts were identified on postoperative CT images and matched to known electrode geometry. Cortical electrodes were projected onto MRI-rendered pial surfaces and adjusted if needed. For group-level reconstructions, electrode locations were normalized into Montreal Neurological Institute (MNI) space and visualized on the FreeSurfer average cortical surface.^45,46^

### Rover wearable devices

To facilitate the collection of at-home kinematic data, we employed lightweight wearable devices (WDs; Sensoplex Inc., Rover; **Fig. 1d**) containing a triaxial gyroscope, accelerometer, and magnetometer (sampling rate [SR]: 100 Hz). These devices were worn around the ankles bilaterally and recorded data on a secure digital (SD) card.

Recordings were uploaded to a secure cloud at regular intervals. Raw WD signals were extracted and formatted with custom code. WD acceleration signals were manually aligned with accelerometry from the Summit RC+S INS to unify neural-kinematic time-domain signals as described previously.^47^

The Rover WDs store gyroscopic information in a quaternion format, which can be represented as follows, avoiding the issue of gimbal lock and ensuring computational efficiency during high-frequency movement:

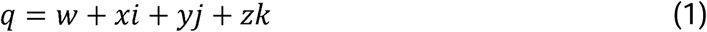

Heading, or yaw, was derived from quaternions by normalizing the quaternion signals before converting to Euler angles (MATLAB *quat2eul* function):

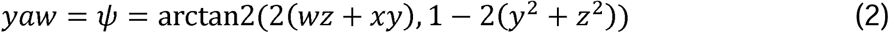

To address discontinuities caused by transitions across the 0°/360° boundary (e.g., a left turn from 90° to 300° causing an arbitrary jump from 0° to 360°), heading signals were unwrapped (MATLAB *unwrap* function). Finally, heading signals were lowpass filtered below 1 Hz using a 4^th^ order Butterworth filter. Angular velocity was calculated as the first derivative of the heading signal. Turns were identified by local peaks in angular velocity above subject-specific thresholds. Valid turns required angular velocity peaks in both left and right leg heading signals to occur within short defined temporal windows to prevent aberrant labeling. Turn direction (i.e., left vs right turns) was determined using the sign of the angular velocity. Periods of straight-walking were defined using periods with low intra-epoch heading variability. Non-walking epochs were excluded from all analyses.

### Validation of gait state labeling

To validate the ability for Rover WDs to accurate distinguish a) turning vs straight-walking epochs and b) left vs right turns for all subjects, we assessed their labeling performance during controlled in-lab trials **(Supplementary Fig. S1)**. During these sessions, subjects performed overground walking loops with left- and right-sided turns. In addition to wearing the Rover WDs, subjects were also equipped with an inertial measurement unit (IMU) system (Xsens, Movella, Netherlands). Each IMU recorded the position, velocity, acceleration, angular velocity, and angular acceleration of the respective body part. Video recordings were captured for each trial.

Ground-truth gait states and turn directions were defined using pelvic angular velocity signals (SR: 60 Hz). A 1.5 Hz lowpass filter was applied to the kinematic signals and patient-specific thresholds were used to define turn starts and stops. All ground-truth labels were manually verified using the video recordings and corrected if necessary. WD-based labels of turning and straight-walking for each 1-second epoch were then compared against known ground-truth behavior. A segment of a sample trial is illustrated in **Fig. 2b**. Accuracy, sensitivity, and specificity of these labels were calculated for each subject across multiple trials; summary metrics for all subjects are presented in **Fig. 2c** and **2d**.

### At-home neural-kinematic data collection and alignment

Subjects streamed at-home multi-site neural data from their Summit RC+S INS (SR: 500 Hz) while wearing Rover WDs for as many hours and across as many days as possible. Subjects remained on clinically optimized stimulation settings and standard PD pharmacological regimens during these recordings. Subjects were not provided with any specific instructions on required recordings times or behavior during recordings, instead allowing them to perform unsupervised daily activities. A summary of recording characteristics is provided in **Supplementary Table S1.** Neural and kinematic signals were synchronized by manual alignment of Summit RC+S and Rover WD acceleration signals using a custom MATLAB graphical user interface (GUI) as previously described.^47^ All recordings were extensively verified to ensure alignment throughout the duration of the session. Recordings were partitioned into 1-second periods and each epoch was labeled as turning or straight-walking based on the method described above.

### Neural data preprocessing

Raw neural time-domain signals were bandpass filtered from 1 to 150 Hz using a 6th order Butterworth filter. ECG and stimulation artifacts were removed using a template subtraction pipeline (https://github.com/lhart1216/PerceptHammer).^48^ All epochs with significant Summit RC+S packet loss or idiosyncratic artifacts were excluded from subsequent analyses.

### Canonical band power and coherence calculation

Time-domain neural signals from each 1-second epoch were converted to the frequency domain using Welch’s method (MATLAB *pwelch* function; 200 ms windows with 90% overlap). Canonical frequency bands were defined as delta (δ; 1-4 Hz), theta (θ; 4-8 Hz), alpha ( ; 8-13 Hz), beta (β; 13-30 Hz), and low gamma (γ_low_; 30-50 Hz). Average power was calculated within each canonical frequency range during each epoch. Power distributions were then compared between turning and straight-walking states. Interregional coherence (i.e., GP-M1, GP-PM, and M1-PM) was calculated for each epoch (Matlab *wcoherence* function). Average coherence was calculated within each canonical frequency range during each epoch and values were compared between turning and straight-walking states.

### Beta burst analysis

To characterize one potential mechanism underlying differences in beta power between gait states, we compared beta burst dynamics during turning and straight-walking epochs. Beta bursts were identified by normalizing the neural signal for each epoch using z-scoring and applying a bandpass filter for the entire beta band, low beta band (13-20 Hz), or high beta band (20.5-30 Hz). All bursts with duration greater than 100 ms and amplitude exceeding the 75^th^ percentile of the signal’s amplitude envelope were selected. The count, duration, and amplitude of all bursts within each epoch were calculated and compared between turning and straight-walking states.

### Time-reversed Granger causality

While coherence offers a metric of undirected interregional connectivity, we sought to also characterize directed information flow during turning and straight-walking epochs. To achieve this, we calculated time-reversed Granger causality (TRGC) using a multivariate vector autoregressive model (VAR) with a model order of 20 (MATLAB MVGC toolbox).^49^ Consistent with prior studies, net Granger scores were established by calculating the difference between seed-to-target and target-to-seed connections (e.g. GP→M1 minus M1→GP).^50^ These values were then corrected by subtracting the net Granger scores obtained from time-reversed data via autocovariance transposition, yielding a final metric isolating true inter-regional interactions.^51,52^ Averaged final TRGC scores within canonical frequency bands were compared to zero to establish the existence of significant causality, and then compared between turning and straight-walking to determine whether directional information flow was modulated by walking state.

### Walking state classification

To assess the relative contributions of canonical frequency band power and coherence values to classification of turning vs straight-walking states, logistic regression models using these metrics were trained and tested independently for each hemisphere. Each epoch contributed 30 features (i.e., 5 canonical frequency band power features and 5 canonical frequency band coherence features from each of 3 regions and 5 canonical frequency band coherence features from each of 3 inter-regional combinations). Monte Carlo cross-validation was used to assess the performance of these models (10 iterations; 70/30 train/test split). Gait states were balanced in the training and testing sets. Mean AUC was reported for each model. To allow for visualization of relative feature weights, mean coefficients for each model were rescaled from −1 to 1.

### Turn direction classification

To assess the possibility of decoding turn direction (i.e., left vs right) using canonical frequency band power and coherence values, logistic regression models using these metrics were also independently trained and tested for each hemisphere. Only turning epochs were used; each epoch again contributed 30 features and Monte Carlo cross-validation was used to assess the performance of these models (10 iterations; 70/30 train/test split). Turn directions were balanced in the training and testing sets. Mean AUC was reported for each model. For models achieving above-chance decoding performance, mean coefficients were rescaled from −1 to 1 and visualized.

### General statistical methods

Differences in power and coherence between turning and straight-walking states were compared within each hemisphere using a two-sided Wilcoxon rank-sum test. To determine if TRGC scores significantly differed from zero, we employed a sign-flip permutation test. A null distribution was generated by randomly inverting the signs of the averaged TRGC values over 1,000 iterations. For comparisons between walking states condition labels were randomly shuffled to construct a surrogate distribution of mean differences. To evaluate the performance of our classification models, trial labels were randomly shuffled, and models were retrained to generate a null distribution of chance AUCs. In all cases, empirical p-values were calculated by comparing observed values to the respective null distributions. To control for the false discovery rate (FDR) associated with multiple testing, p-values were corrected using the Benjamini-Hochberg method.^53^ Significance was assessed at an FDR-adjusted p<0.05.

## Data Availability

All data produced in the present work are contained in the manuscript and associated supplementary material.

## ACKNOWLEDGEMENTS

We thank Lauren Hammer for artifact removal code (https://github.com/lhart1216/PerceptHammer).

