## Supplementary Material for "Neural signatures of real-world turning during naturalistic locomotion in Parkinson’s Disease"

**Figure S1. Validation of Rover wearable device walking state labeling……………………………..…….2**

**Figure S2. Average power spectral densities by walking state……………………………………..………3**

**Figure S3. Average coherence spectra by walking state……………………………………………….……4**

**Figure S4. Summary of Granger causality by walking state and recording setting………………...……5**

**Table S1. Summary of at-home neural-kinematic recordings……………………………….……………...7**

**Table S2. Comparison of canonical frequency band power between walking states…...……….……..8**

**Table S3. Comparison of beta burst dynamics between walking states………………………..………...9**

**Table S4. Comparison of canonical band coherence between walking states……………………...….10**

**Table S5. Summary of logistic regression models for classifying gait state…………….………..…….11**

**Table S6. Comparison of TRGC scores between in-clinic and at-home recordings………….....…….12**

**Table S7. Summary of logistic regression models for classifying turn direction…….………..……….13**

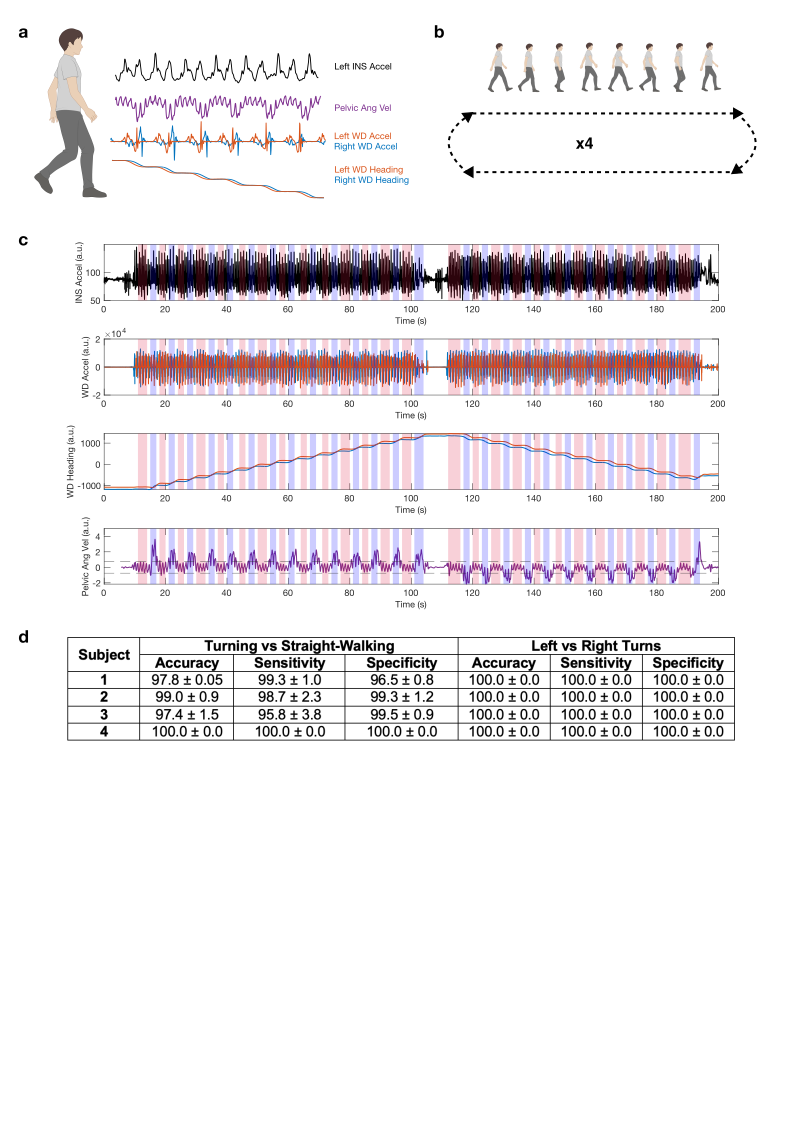

**Figure S1. Validation of Rover wearable device walking state labeling | a**, Sample signals are shown from unilateral INS, pelvic IMU, and bilateral WDs during an in-laboratory walking state labeling validation session. **b,** During in-laboratory trials, subjects performed overground walking loops with left- and right-sided turns. **c,** Aligned INS acceleration, WD acceleration, WD heading, and pelvic IMU angular velocity signals are shown from a 200-second in-laboratory validation session. Blue patches indicate periods labeled as turning based on WD heading signals whereas red patches indicate straight-walking labels. **d,** Accuracy, sensitivity, and specificity are shown for WD-based labeling of turning vs straight-walking epochs and left vs right turns during in-laboratory validation trials. Metrics are shown as mean ± SD. IMU, inertial measurement unit; INS, implantable neurostimulator; WD, wearable device; SD, standard deviation.

**Figure S2. Average power spectral densities by walking state |** Mean power spectral densities are shown from 0 to 50 Hz for turning (blue) and straight walking (red) epochs from all hemispheres. GP, globus pallidus; M1, primary motor cortex; PM, premotor cortex.

**Figure S3. Average coherence spectra by walking state |** Mean coherence spectra are shown from 0 to 50 Hz for turning (blue) and straight walking (red) epochs from all hemispheres. GP, globus pallidus; M1, primary motor cortex; PM, premotor cortex.

**Figure S4. Summary of Granger causality by walking state and recording setting | a,** Mean time-reversed Granger causality scores are shown from 0 to 50 Hz for turning (blue) and straight walking (red) epochs for all cortical-pallidal and cortical-cortical connections in all hemispheres. Shaded regions represent standard error of the mean. **b,** Summary heatmaps show the number of significant hemispheres for each connection and frequency band combination during (left) turning vs a null distribution, (middle) straight walking vs a null distribution, and (right) straight walking vs turning. **c,** Summary heatmaps show the number of significant hemispheres for each connection and frequency band combination during (left) turning during clinic vs home recordings, and (right) straight walking during clinic vs home recordings. TRGC = time-reversed Granger causality; GP, globus pallidus; M1, primary motor cortex; PM, premotor cortex.

| **Subject** | **Hemisphere** | **Number of Recording Days** | **Number of Recording Sessions** | **Total Recording Duration (s)** | **Turning Epochs (s)** | **Straight Walking Epochs (s)** |
| --- | --- | --- | --- | --- | --- | --- |
| **1** | L | 6 | 7 | 36,555 | 1,670 | 6,515 |
| **2** | L | 5 | 5 | 39,584 | 715 | 1,159 |
|  | R | 5 | 5 | 39,584 | 715 | 1,159 |
| **3** | L | 8 | 8 | 5,488 | 498 | 927 |
|  | R | 8 | 8 | 5,488 | 498 | 927 |
| **4** | L | 12 | 24 | 85,790 | 1,503 | 9,273 |

**Supplementary Table S1. Summary of at-home neural-kinematic recordings |** Details of all at-home neural-kinematic recordings performed by subjects are shown, including the number of individual days during which recordings were performed, the number of individual recording sessions, total recorded duration, and total analyzed duration of turning and straight-walking epochs. L, left; R, right.

| **Subject** | **Region** | **Median Power within Canonical Frequency Band (dB/Hz)** | | | | | | | | | | | | | | |
| --- | --- | --- | --- | --- | --- | --- | --- | --- | --- | --- | --- | --- | --- | --- | --- | --- |
|  |  | **δ (1-4 Hz)** | | | **Θ (4-8 Hz)** | | | **ɑ (8-13 Hz)** | | | **β (13-30 Hz)** | | | **γ_low_ (30-50 Hz)** | | |
|  |  | **SW** | **T** | **p** | **SW** | **T** | **p** | **SW** | **T** | **p** | **SW** | **T** | **p** | **SW** | **T** | **p** |
| **1** | **L GP** | -58.0 | -59.7 | 1.14e-79 | -60.1 | -62.0 | 7.22e-133 | -63.2 | -64.8 | 1.13e-108 | -69.1 | -69.8 | 7.30e-62 | -75.9 | -75.9 | 0.826 |
|  | **L M1** | -49.9 | -51.1 | 1.24e-52 | -50.6 | -51.9 | 4.65e-64 | -51.6 | -52.7 | 1.06e-29 | -50.9 | -51.6 | 6.57e-5 | -54.6 | -54.2 | 3.14e-13 |
|  | **L PM** | -48.9 | -50.2 | 1.37e-30 | -47.0 | -48.8 | 5.15e-38 | -41.9 | -45.4 | 1.54e-42 | **-42.6** | **-43.6** | **1.54e-13** | -49.9 | -50.2 | 0.0013 |
| **2** | **L GP** | -56.6 | -56.6 | 0.324 | -60.0 | -59.2 | 5.45e-8 | -63.1 | -62.1 | 3.16e-14 | -66.9 | -66.1 | 3.55e-11 | -74.2 | -74.0 | 0.017 |
|  | **L M1** | -55.8 | -56.0 | 0.163 | -55.9 | -56.2 | 0.060 | -59.0 | -59.4 | 0.002 | -60.9 | -62.2 | 5.5e-13 | -64.5 | -64.9 | 3.94e-6 |
|  | **L PM** | -55.3 | -55.0 | 0.010 | -55.4 | -54.7 | 4.65e-5 | -59.4 | -58.7 | 8.65e-6 | **-62.7** | **-63.4** | **3.18e-6** | -69.1 | -69.4 | 1.87e-6 |
|  | **R GP** | -54.2 | -53.3 | 2.67e-9 | -57.5 | -56.9 | 1.35e-7 | -62.0 | -61.1 | 8.17e-13 | -68.0 | -67.6 | 1.90e-6 | -73.9 | -73.8 | 0.354 |
|  | **R M1** | -55.8 | -54.8 | 8.03e-9 | -56.3 | -55.5 | 3.50e-6 | -59.9 | -58.7 | 1.01e-7 | -61.7 | -61.1 | 0.263 | -65.8 | -64.9 | 1.26e-9 |
|  | **R PM** | -59.0 | -58.1 | 1.61e-9 | -58.7 | -58.3 | 4.87e-4 | -62.1 | -61.6 | 1.64e-4 | **-64.3** | **-64.8** | **3.02e-5** | -70.2 | -70.0 | 0.052 |
| **3** | **L GP** | -42.1 | -42.3 | 0.299 | -40.5 | -40.7 | 0.105 | -39.2 | -39.2 | 0.763 | -43.6 | -43.5 | 0.253 | -56.1 | -56.0 | 0.868 |
|  | **L M1** | -56.2 | -56.9 | 1.35e-5 | -56.5 | -57.0 | 2.55e-4 | -57.8 | -58.4 | 0.007 | -59.2 | -59.6 | 0.008 | -65.8 | -65.8 | 0.736 |
|  | **L PM** | -55.3 | -56.0 | 2.96e-5 | -54.9 | -55.3 | 0.003 | -55.9 | -56.4 | 0.019 | **-56.3** | **-57.0** | **1.85e-5** | -64.5 | -64.9 | 0.006 |
|  | **R GP** | -47.0 | -47.4 | 0.067 | -47.3 | -47.6 | 0.031 | -48.1 | -48.1 | 0.214 | -49.8 | -49.9 | 0.460 | -58.6 | -58.5 | 0.495 |
|  | **R M1** | -59.8 | -59.8 | 0.226 | -59.8 | -60.3 | 4.28e-4 | -60.7 | -61.1 | 0.012 | -60.9 | -61.2 | 0.043 | -68.1 | -68.1 | 0.611 |
|  | **R PM** | -57.7 | -57.9 | 0.295 | -56.7 | -57.1 | 0.020 | -57.8 | -58.1 | 0.020 | **-57.6** | **-58.3** | **0.002** | -65.7 | -65.4 | 0.002 |
| **4** | **L GP** | -36.6 | -36.8 | 1.58e-9 | -34.8 | -34.9 | 3.54e-4 | -35.4 | -35.4 | 0.143 | -42.6 | -42.7 | 0.624 | -53.8 | -53.8 | 0.934 |
|  | **L M1** | -54.8 | -56.3 | 1.07e-84 | -54.8 | -56.5 | 2.51e-110 | -57.2 | -58.8 | 3.77e-83 | -58.3 | -59.2 | 3.95e-14 | -64.5 | -64.4 | 0.011 |
|  | **L PM** | -54.0 | -54.5 | 1.97e-11 | -52.8 | -53.3 | 8.78e-12 | -55.7 | -55.9 | 1.30e-4 | **-57.5** | **-58.4** | **6.63e-31** | -64.4 | -64.3 | 1.60e-4 |

**Supplementary Table S2. Comparison of canonical frequency band power between turning and straight-walking epochs |** Median pallidal and cortical power values are shown across delta (δ), theta (Θ), alpha (ɑ), beta (β), and low gamma (γlow) canonical frequency bands. Power differences between straight-walking (SW) and turning (T) epochs were assessed using two-sided Wilcoxon rank sum tests with Benjamini-Hochberg correction for multiple comparisons. Red shading indicates comparison with higher power during straight-walking. Blue shading indicates comparisons with higher power during turning. Gray shading indicates comparisons without a significant difference. Premotor cortex beta power was lower during turning epochs compared to straight-walking epochs in all hemispheres, indicated by hatched boxes. Uncorrected p-values are shown. L, left; R, right; GP, globus pallidus; M1, primary motor cortex; PM, premotor cortex.

**a.**

| **Region** | **Range** | **Subject 1** | | | **Subject 2** | | | | | | **Subject 3** | | | | | | **Subject 4** | | |
| --- | --- | --- | --- | --- | --- | --- | --- | --- | --- | --- | --- | --- | --- | --- | --- | --- | --- | --- | --- |
|  |  | **L** | | | **L** | | | **R** | | | **L** | | | **R** | | | **L** | | |
|  |  | **SW** | **T** | **p** | **SW** | **T** | **p** | **SW** | **T** | **p** | **SW** | **T** | **p** | **SW** | **T** | **p** | **SW** | **T** | **p** |
| **GP** | **β** | 1 | 0 | 2.43e-32 | 1 | 1 | 0.001 | 0 | 1 | 2.98e-18 | 1 | 1 | 1.86e-5 | 1 | 1 | 1.66e-4 | 2 | 1 | 2.51e-11 |
|  | **β_low_** | 1 | 1 | 1.71e-49 | 1 | 2 | 5.73e-24 | 1 | 1 | 1.83e-6 | 1 | 1 | 2.13e-5 | 1 | 1 | 4.81e-6 | 2 | 2 | 3.70e-11 |
|  | **β_high_** | 1 | 1 | 3.48e-19 | 1 | 2 | 2.04e-25 | 1 | 1 | 5.33e-7 | 1 | 1 | 8.65e-6 | 1 | 1 | 1.13e-4 | 2 | 1 | 4.01e-6 |
| **M1** | **β** | 1 | 1 | 0.694 | 0 | 0 | 0.814 | 0 | 0 | 0.812 | 1 | 0 | 1.82e-7 | 0 | 0 | 0.002 | 0 | 0 | 0.056 |
|  | **β_low_** | 1 | 1 | 4.61e-11 | 0 | 1 | 8.29e-5 | 0 | 1 | 1.64e-13 | 1 | 0 | 2.45e-9 | 1 | 1 | 8.46e-9 | 1 | 0 | 5.86e-17 |
|  | **β_high_** | 1 | 1 | 0.310 | 0 | 0 | 0.756 | 0 | 1 | 6.62e-5 | 1 | 0 | 2.61e-5 | 1 | 1 | 0.052 | 1 | 1 | 0.728 |
| **PM** | **β** | 1 | 1 | 8.28e-6 | 0 | 0 | 0.361 | 0 | 0 | 0.010 | 1 | 0 | 1.12e-5 | 0 | 0 | 0.038 | 0 | 0 | 3.99e-19 |
|  | **β_low_** | 1 | 1 | 9.41e-31 | 0 | 1 | 3.19e-13 | 0 | 1 | 9.19e-12 | 1 | 1 | 1.36e-5 | 1 | 1 | 2.45e-6 | 1 | 1 | 8.86e-6 |
|  | **β_high_** | 1 | 1 | 7.82e-4 | 0 | 0 | 0.288 | 0 | 1 | 0.343 | 1 | 1 | 0.003 | 1 | 0 | 1.32e-4 | 1 | 1 | 7.13e-27 |

**b.**

| **Region** | **Range** | **Subject 1** | | | **Subject 2** | | | | | | **Subject 3** | | | | | | **Subject 4** | | |
| --- | --- | --- | --- | --- | --- | --- | --- | --- | --- | --- | --- | --- | --- | --- | --- | --- | --- | --- | --- |
|  |  | **L** | | | **L** | | | **R** | | | **L** | | | **R** | | | **L** | | |
|  |  | **SW** | **T** | **p** | **SW** | **T** | **p** | **SW** | **T** | **p** | **SW** | **T** | **p** | **SW** | **T** | **p** | **SW** | **T** | **p** |
| **GP** | **β** | 0.006 | 0.006 | 6.23e-6 | 0.327 | 0.347 | 1.25e-20 | 0.075 | 0.007 | 1.25e-20 | 0.194 | 0.199 | 0.002 | 0.090 | 0.090 | 0.320 | 0.213 | 0.213 | 0.078 |
|  | **β_low_** | 0.004 | 0.004 | 1.26e-11 | 0.165 | 0.169 | 1.26e-7 | 0.042 | 0.004 | 1.26e-7 | 0.113 | 0.114 | 0.026 | 0.047 | 0.047 | 0.686 | 0.134 | 0.135 | 0.001 |
|  | **β_high_** | 0.003 | 0.003 | 0.053 | 0.166 | 0.177 | 3.12e-27 | 0.035 | 0.004 | 3.12e-27 | 0.085 | 0.087 | 0.002 | 0.048 | 0.049 | 0.487 | 0.082 | 0.082 | 0.024 |
| **M1** | **β** | 0.071 | 0.080 | 4.05e-14 | 0.020 | 0.022 | 0.003 | 0.018 | 0.023 | 0.003 | 0.022 | 0.023 | 0.767 | 0.017 | 0.017 | 0.969 | 0.028 | 0.031 | 1.50e-14 |
|  | **β_low_** | 0.037 | 0.041 | 1.04e-6 | 0.008 | 0.008 | 0.219 | 0.007 | 0.012 | 0.219 | 0.012 | 0.011 | 0.112 | 0.009 | 0.009 | 0.749 | 0.012 | 0.012 | 0.603 |
|  | **β_high_** | 0.040 | 0.045 | 2.37e-26 | 0.013 | 0.013 | 0.109 | 0.013 | 0.015 | 0.109 | 0.014 | 0.014 | 0.105 | 0.011 | 0.011 | 0.791 | 0.019 | 0.020 | 1.56e-5 |
| **PM** | **β** | 0.148 | 0.151 | 0.003 | 0.015 | 0.015 | 0.390 | 0.012 | 0.013 | 0.390 | 0.030 | 0.029 | 0.001 | 0.026 | 0.028 | 0.001 | 0.026 | 0.025 | 0.046 |
|  | **β_low_** | 0.101 | 0.103 | 4.60e-4 | 0.008 | 0.008 | 0.142 | 0.006 | 0.007 | 0.142 | 0.017 | 0.016 | 0.001 | 0.013 | 0.014 | 2.45e-6 | 0.013 | 0.012 | 4.86e-10 |
|  | **β_high_** | 0.078 | 0.080 | 2.20e-5 | 0.009 | 0.008 | 6.07e-7 | 0.008 | 0.008 | 6.07e-7 | 0.018 | 0.018 | 0.014 | 0.018 | 0.019 | 1.32e-4 | 0.017 | 0.017 | 1.01e-5 |

**c.**

| **Region** | **Range** | **Subject 1** | | | **Subject 2** | | | | | | **Subject 3** | | | | | | **Subject 4** | | |
| --- | --- | --- | --- | --- | --- | --- | --- | --- | --- | --- | --- | --- | --- | --- | --- | --- | --- | --- | --- |
|  |  | **L** | | | **L** | | | **R** | | | **L** | | | **R** | | | **L** | | |
|  |  | **SW** | **T** | **p** | **SW** | **T** | **p** | **SW** | **T** | **p** | **SW** | **T** | **p** | **SW** | **T** | **p** | **SW** | **T** | **p** |
| **GP** | **β** | 0.130 | 0.126 | 0.049 | 0.110 | 0.106 | 1.15e-10 | 0.114 | 0.110 | 0.001 | 0.154 | 0.156 | 0.355 | 0.116 | 0.118 | 0.246 | 0.154 | 0.150 | 0.001 |
|  | **β_low_** | 0.160 | 0.152 | 1.51e-6 | 0.147 | 0.150 | 3.35e-8 | 0.144 | 0.160 | 3.95e-16 | 0.176 | 0.178 | 0.001 | 0.162 | 0.162 | 0.873 | 0.186 | 0.184 | 0.006 |
|  | **β_high_** | 0.132 | 0.130 | 0.023 | 0.136 | 0.134 | 0.065 | 0.122 | 0.142 | 2.16e-22 | 0.152 | 0.154 | 2.01e-4 | 0.146 | 0.146 | 0.848 | 0.140 | 0.138 | 0.005 |
| **M1** | **β** | 0.144 | 0.160 | 1.24e-15 | 0.160 | 0.162 | 0.823 | 0.150 | 0.142 | 0.004 | 0.144 | 0.150 | 0.895 | 0.136 | 0.131 | 0.039 | 0.144 | 0.160 | 5.00e-11 |
|  | **β_low_** | 0.188 | 0.206 | 4.18e-9 | 0.158 | 0.162 | 0.163 | 0.164 | 0.180 | 1.23e-4 | 0.164 | 0.162 | 0.318 | 0.154 | 0.154 | 0.358 | 0.154 | 0.158 | 0.016 |
|  | **β_high_** | 0.142 | 0.152 | 1.24e-11 | 0.168 | 0.148 | 0.005 | 0.156 | 0.132 | 1.49e-8 | 0.158 | 0.158 | 0.331 | 0.150 | 0.136 | 0.006 | 0.158 | 0.164 | 0.002 |
| **PM** | **β** | 0.144 | 0.146 | 0.129 | 0.138 | 0.140 | 1.000 | 0.140 | 0.130 | 0.006 | 0.155 | 0.142 | 0.041 | 0.151 | 0.156 | 0.205 | 0.142 | 0.138 | 0.124 |
|  | **β_low_** | 0.196 | 0.202 | 4.35e-4 | 0.162 | 0.162 | 0.840 | 0.160 | 0.158 | 0.060 | 0.176 | 0.170 | 0.069 | 0.160 | 0.168 | 0.172 | 0.154 | 0.152 | 0.320 |
|  | **β_high_** | 0.144 | 0.148 | 0.003 | 0.148 | 0.140 | 0.009 | 0.144 | 0.134 | 3.65e-5 | 0.142 | 0.144 | 0.895 | 0.160 | 0.169 | 0.041 | 0.148 | 0.142 | 0.082 |

**Supplementary Table S3. Comparison of beta burst dynamics between turning and straight-walking epochs |** Median beta burst **(a)** count per epoch, **(b)** amplitude, and **(c)** duration are shown across beta (β, 13-30 Hz), low beta (β_low_, 13-20 Hz), and high beta (β_high_, 13-20 Hz) frequency bands. Differences in burst count, amplitude, and duration between straight-walking (SW) and turning (T) epochs were assessed using two-sided Wilcoxon rank sum tests with Benjamini-Hochberg correction for multiple comparisons. Uncorrected p-values are shown. Red shading indicates comparison with higher values during straight-walking. Blue shading indicates comparisons with higher values during turning. Gray shading indicates comparisons without a significant difference. L, left; R, right; GP, globus pallidus; M1, primary motor cortex; PM, premotor cortex.

| **Subject** | **Region** | **Median Coherence within Canonical Frequency Band** | | | | | | | | | | | | | | |
| --- | --- | --- | --- | --- | --- | --- | --- | --- | --- | --- | --- | --- | --- | --- | --- | --- |
|  |  | **δ (1-4 Hz)** | | | **Θ (4-8 Hz)** | | | **ɑ (8-13 Hz)** | | | **β (13-30 Hz)** | | | **γ_low_ (30-50 Hz)** | | |
|  |  | **SW** | **T** | **p** | **SW** | **T** | **p** | **SW** | **T** | **p** | **SW** | **T** | **p** | **SW** | **T** | **p** |
| **1** | **L GP-M1** | 0.304 | 0.305 | 0.275 | 0.229 | 0.221 | 0.004 | 0.230 | 0.234 | 0.318 | 0.268 | 0.257 | 2.14e-7 | 0.262 | 0.259 | 0.002 |
|  | **L GP-PM** | 0.279 | 0.287 | 0.534 | 0.204 | 0.206 | 0.072 | 0.199 | 0.213 | 7.39e-10 | 0.259 | 0.256 | 0.032 | 0.268 | 0.264 | 0.031 |
|  | **L M1-PM** | 0.312 | 0.325 | 0.168 | 0.232 | 0.243 | 4.67e-4 | 0.281 | 0.279 | 0.067 | 0.294 | 0.295 | 0.842 | 0.282 | 0.292 | 2.33e-9 |
| **2** | **L GP-M1** | 0.277 | 0.272 | 0.680 | 0.229 | 0.231 | 0.628 | 0.244 | 0.235 | 0.327 | 0.251 | 0.253 | 0.656 | 0.267 | 0.270 | 0.221 |
|  | **L GP-PM** | 0.279 | 0.265 | 0.304 | 0.219 | 0.234 | 0.204 | 0.224 | 0.228 | 0.674 | 0.253 | 0.250 | 0.585 | 0.266 | 0.262 | 0.103 |
|  | **L M1-PM** | 0.291 | 0.298 | 0.857 | 0.259 | 0.256 | 0.479 | 0.266 | 0.263 | 0.573 | 0.312 | 0.280 | 3.99e-12 | 0.327 | 0.309 | 8.45e-6 |
|  | **R GP-M1** | 0.291 | 0.363 | 1.49e-4 | 0.225 | 0.287 | 4.78e-12 | 0.240 | 0.282 | 2.48e-12 | 0.243 | 0.270 | 1.23e-14 | 0.271 | 0.283 | 1.58e-5 |
|  | **R GP-PM** | 0.296 | 0.344 | 3.19e-4 | 0.239 | 0.265 | 1.03e-4 | 0.247 | 0.276 | 1.27e-6 | 0.251 | 0.272 | 2.86e-9 | 0.275 | 0.284 | 8.04e-4 |
|  | **R M1-PM** | 0.329 | 0.335 | 0.175 | 0.298 | 0.300 | 0.618 | 0.296 | 0.325 | 5.01e-4 | 0.367 | 0.358 | 0.046 | 0.370 | 0.360 | 0.052 |
| **3** | **L GP-M1** | 0.277 | 0.272 | 0.878 | 0.203 | 0.202 | 0.890 | 0.187 | 0.185 | 0.717 | 0.194 | 0.192 | 0.625 | 0.220 | 0.218 | 0.344 |
|  | **L GP-PM** | 0.251 | 0.267 | 0.147 | 0.208 | 0.201 | 0.951 | 0.187 | 0.182 | 0.288 | 0.192 | 0.188 | 0.588 | 0.227 | 0.227 | 0.818 |
|  | **L M1-PM** | 0.300 | 0.294 | 0.730 | 0.250 | 0.247 | 0.756 | 0.278 | 0.277 | 0.284 | 0.342 | 0.329 | 0.015 | 0.280 | 0.281 | 0.700 |
|  | **R GP-M1** | 0.296 | 0.268 | 0.305 | 0.211 | 0.210 | 0.777 | 0.223 | 0.220 | 0.701 | 0.219 | 0.218 | 0.881 | 0.232 | 0.235 | 0.326 |
|  | **R GP-PM** | 0.273 | 0.274 | 0.546 | 0.212 | 0.208 | 0.499 | 0.221 | 0.219 | 0.680 | 0.212 | 0.215 | 0.365 | 0.233 | 0.235 | 0.494 |
|  | **R M1-PM** | 0.307 | 0.286 | 0.292 | 0.251 | 0.246 | 0.525 | 0.278 | 0.270 | 0.154 | 0.392 | 0.379 | 0.016 | 0.321 | 0.325 | 0.439 |
| **4** | **L GP-M1** | 0.295 | 0.299 | 0.636 | 0.225 | 0.215 | 0.020 | 0.201 | 0.197 | 0.082 | 0.182 | 0.185 | 0.585 | 0.223 | 0.223 | 0.290 |
|  | **L GP-PM** | 0.287 | 0.279 | 0.365 | 0.226 | 0.218 | 0.031 | 0.204 | 0.196 | 0.055 | 0.185 | 0.189 | 0.278 | 0.220 | 0.220 | 0.553 |
|  | **L M1-PM** | 0.317 | 0.312 | 0.894 | 0.269 | 0.262 | 0.022 | 0.264 | 0.256 | 0.002 | 0.355 | 0.323 | 2.45e-20 | 0.337 | 0.331 | 0.006 |

**Supplementary Table S4. Comparison of canonical band coherence between gait states |** Median cortical-pallidal and cortical-cortical coherence values are shown across delta (δ), theta (Θ), alpha (ɑ), beta (β), and low gamma (γlow) canonical frequency bands. Power differences between straight-walking (SW) and turning (T) epochs were assessed using two-sided Wilcoxon rank sum tests with Benjamini-Hochberg correction for multiple comparisons. Uncorrected p-values are shown. Red shading indicates comparison with higher coherence during straight-walking. Blue shading indicates comparisons with higher coherence during turning. Gray shading indicates comparisons without a significant difference. L, left; R, right; GP, globus pallidus; M1, primary motor cortex; PM, premotor cortex.

| **Value** | **Region(s)** | **Frequency Band** | **Subject** | | | | | |
| --- | --- | --- | --- | --- | --- | --- | --- | --- |
|  |  |  | **1** | **2** | | **3** | | **4** |
|  |  |  | **L** | **L** | **R** | **L** | **R** | **L** |
| **Power** | **GP** | **δ** | 0.24 | -0.40 | 0.63 | -0.08 | -0.27 | -0.54 |
|  |  | **Θ** | -0.51 | 0.02 | -0.44 | -0.45 | -0.83 | 0.63 |
|  |  | **ɑ** | -0.07 | 0.29 | 1 | -0.15 | 0.04 | -0.15 |
|  |  | **β** | 0.01 | 0.25 | 0.63 | 1 | 0.20 | 0.11 |
|  |  | **γ_low_** | 1 | -0.35 | -0.70 | -0.11 | 0.01 | 0.24 |
|  | **M1** | **δ** | 0.11 | -0.26 | 0.42 | -0.83 | -0.09 | -0.93 |
|  |  | **Θ** | -0.12 | -0.47 | -0.32 | 0.21 | -0.71 | -0.52 |
|  |  | **ɑ** | 0.35 | 0.17 | 0.72 | 0.04 | -0.15 | -0.87 |
|  |  | **β** | 0.79 | -0.59 | 0.06 | -0.12 | -0.17 | 1 |
|  |  | **γ_low_** | 0.90 | -0.13 | 0.78 | 0.12 | -0.31 | 0.81 |
|  | **PM** | **δ** | 0.08 | 0.08 | 0.62 | -0.72 | 0.08 | 0.24 |
|  |  | **Θ** | 0.69 | -0.67 | -0.4 | -0.10 | -0.42 | -0.55 |
|  |  | **ɑ** | -1 | 1 | 0.80 | 0.52 | 0.26 | 0.79 |
|  |  | **β** | 0.19 | -1 | -1 | -1 | -1 | -1 |
|  |  | **γ_low_** | 0.67 | -0.61 | 0.33 | -0.09 | 1 | 0.69 |
| **Coherence** | **GP-M1** | **δ** | 0.39 | -0.19 | 0.18 | 0.09 | -0.33 | 0.17 |
|  |  | **Θ** | 0.33 | -0.08 | 0.39 | -0.01 | -0.11 | 0.01 |
|  |  | **ɑ** | 0.38 | -0.28 | 0.24 | 0.01 | -0.09 | 0.11 |
|  |  | **β** | 0.10 | 0.06 | 0.45 | 0.06 | -0.39 | 0.13 |
|  |  | **γ_low_** | 0.06 | 0.07 | 0.37 | -0.25 | 0.05 | 0.06 |
|  | **GP-PM** | **δ** | 0.41 | -0.29 | 0.31 | 0.25 | 0.06 | 0.10 |
|  |  | **Θ** | 0.31 | -0.02 | 0.18 | 0.02 | -0.21 | 0.06 |
|  |  | **ɑ** | 0.40 | -0.15 | 0.2 | -0.29 | -0.27 | -0.19 |
|  |  | **β** | 0.25 | -0.13 | 0.45 | 0.04 | 0.15 | 0.18 |
|  |  | **γ_low_** | 0.08 | -0.23 | 0.09 | 0.01 | -0.14 | 0.09 |
|  | **M1-PM** | **δ** | 0.26 | -0.05 | 0.23 | -0.03 | -0.38 | 0.31 |
|  |  | **Θ** | 0.52 | -0.12 | 0.16 | 0.44 | -0.03 | 0.01 |
|  |  | **ɑ** | 0.25 | 0.09 | 0.28 | -0.12 | -0.09 | 0.22 |
|  |  | **β** | 0.26 | -0.78 | 0.13 | -0.43 | -0.57 | -0.55 |
|  |  | **γ_low_** | 0.44 | -0.01 | 0.11 | 0.13 | 0.30 | 0.19 |
| **AUC** | | | 0.77 | 0.69 | 0.73 | 0.58 | 0.56 | 0.73 |
| **p** | | | <0.001 | <0.001 | <0.001 | 0.018 | 0.048 | <0.001 |

**Supplementary Table S5. Summary of logistic regression models for classifying gait states |** Logistic regression models were trained to classify turning vs straight-walking using average power and coherence within canonical frequency bands from GP, M1, and PM signals, separately for each hemisphere. Model coefficients are shown, rescaled from -1 to 1. Mean AUC is reported for each model, with 10-fold cross validation. Empirical one-sided p-values were calculated by randomly shuffling true labels 1,000 times and determining the AUC for each permutation. L, left; R, right; GP, globus pallidus; M1, primary motor cortex; PM, premotor cortex.

| **Subject** | **Region** | **Mean Granger Score within Canonical Frequency Band During Turning** | | | | | | | | | | | | | | |
| --- | --- | --- | --- | --- | --- | --- | --- | --- | --- | --- | --- | --- | --- | --- | --- | --- |
|  |  | **δ (1-4 Hz)** | | | **Θ (4-8 Hz)** | | | **ɑ (8-13 Hz)** | | | **β (13-30 Hz)** | | | **γ_low_ (30-50 Hz)** | | |
|  |  | **Clinic** | **Home** | **p** | **Clinic** | **Home** | **p** | **Clinic** | **Home** | **p** | **Clinic** | **Home** | **p** | **Clinic** | **Home** | **p** |
| **1** | **L GP-M1** | 0.018 | 0.002 | 0.58 | 0.012 | -0.003 | 0.460 | -0.006 | -0.009 | 0.839 | -0.042 | -0.043 | 0.978 | -0.014 | -0.012 | 0.882 |
|  | **L GP-PM** | 0.008 | -0.007 | 0.413 | 0.005 | -0.005 | 0.479 | 0.001 | -0.001 | 0.928 | -0.014 | -0.004 | 0.429 | -0.015 | -0.003 | 0.254 |
|  | **L M1-PM** | 0.059 | 0.062 | 0.939 | 0.05 | 0.065 | 0.566 | 0.06 | 0.089 | 0.34 | 0.076 | 0.046 | 0.179 | 0.052 | 0.015 | 0.011 |
| **2** | **L GP-M1** | -0.005 | 0.001 | 0.868 | -0.016 | -0.004 | 0.583 | -0.02 | -0.009 | 0.555 | 0.001 | -0.045 | 0.018 | -0.004 | -0.023 | 0.113 |
|  | **L GP-PM** | 0.042 | 0.009 | 0.286 | 0.019 | 0.002 | 0.454 | 0.003 | -0.008 | 0.578 | -0.032 | -0.017 | 0.300 | -0.024 | -0.008 | 0.195 |
|  | **L M1-PM** | 0.031 | 0.04 | 0.793 | 0.039 | 0.032 | 0.805 | 0.042 | 0.019 | 0.286 | 0.039 | 0.007 | 0.071 | 0.015 | 0.017 | 0.899 |
|  | **R GP-M1** | -0.041 | 0.113 | 0.011 | 0.302 | 0.008 | <0.001 | 0.175 | -0.03 | <0.001 | -0.045 | -0.044 | 0.962 | -0.146 | -0.057 | <0.001 |
|  | **R GP-PM** | -0.253 | -0.189 | 0.369 | -0.135 | -0.177 | 0.500 | -0.094 | -0.177 | 0.169 | -0.088 | -0.115 | 0.449 | -0.028 | -0.055 | 0.184 |
|  | **R M1-PM** | 0.113 | 0.442 | <0.001 | 0.017 | 0.275 | <0.001 | -0.012 | 0.18 | <0.001 | 0.007 | 0.116 | <0.001 | -0.035 | 0.035 | <0.001 |
| **3** | **L GP-M1** | 0.024 | -0.008 | 0.275 | 0.026 | -0.009 | 0.087 | 0.025 | -0.011 | 0.080 | -0.012 | -0.010 | 0.890 | 0.003 | -0.004 | 0.559 |
|  | **L GP-PM** | 0.012 | 0.006 | 0.834 | -0.004 | 0.001 | 0.824 | -0.018 | -0.002 | 0.494 | -0.022 | 0.001 | 0.202 | -0.03 | -0.01 | 0.142 |
|  | **L M1-PM** | -0.033 | 0.015 | 0.102 | -0.021 | 0.019 | 0.038 | -0.007 | 0.021 | 0.158 | -0.005 | 0.014 | 0.406 | -0.008 | -0.001 | 0.595 |
|  | **R GP-M1** | -0.03 | 0.023 | 0.076 | -0.028 | 0.014 | 0.045 | -0.031 | 0.008 | 0.049 | -0.018 | -0.004 | 0.434 | -0.019 | -0.005 | 0.206 |
|  | **R GP-PM** | -0.025 | -0.004 | 0.487 | -0.016 | 0.002 | 0.415 | -0.011 | 0.007 | 0.352 | -0.008 | 0.001 | 0.549 | -0.024 | 0.008 | 0.006 |
|  | **R M1-PM** | 0.03 | -0.004 | 0.336 | 0.027 | 0.003 | 0.335 | 0.022 | 0.018 | 0.871 | -0.030 | 0.050 | 0.002 | -0.008 | 0.001 | 0.587 |
| **4** | **L GP-M1** | -0.001 | 0.001 | 0.922 | -0.006 | 0.001 | 0.716 | -0.005 | 0.001 | 0.766 | -0.004 | -0.008 | 0.770 | -0.006 | -0.009 | 0.784 |
|  | **L GP-PM** | -0.022 | -0.004 | 0.409 | -0.022 | -0.011 | 0.581 | -0.013 | -0.013 | 0.999 | 0.010 | -0.010 | 0.123 | -0.002 | -0.004 | 0.796 |
|  | **L M1-PM** | 0.006 | 0.007 | 0.966 | 0.001 | 0.002 | 0.971 | -0.004 | 0.003 | 0.695 | 0.028 | 0.020 | 0.699 | -0.02 | 0.005 | 0.040 |

**a**

**b**

| **Subject** | **Region** | **Mean Granger Score within Canonical Frequency Band During Straight-Walking** | | | | | | | | | | | | | | |
| --- | --- | --- | --- | --- | --- | --- | --- | --- | --- | --- | --- | --- | --- | --- | --- | --- |
|  |  | **δ (1-4 Hz)** | | | **Θ (4-8 Hz)** | | | **ɑ (8-13 Hz)** | | | **β (13-30 Hz)** | | | **γ_low_ (30-50 Hz)** | | |
|  |  | **Clinic** | **Home** | **p** | **Clinic** | **Home** | **p** | **Clinic** | **Home** | **p** | **Clinic** | **Home** | **p** | **Clinic** | **Home** | **p** |
| **1** | **L GP-M1** | 0.028 | 0.009 | 0.391 | 0.025 | -0.004 | 0.076 | 0.018 | -0.019 | 0.007 | -0.005 | -0.066 | <0.001 | -0.001 | -0.023 | 0.009 |
|  | **L GP-PM** | -0.017 | -0.003 | 0.296 | -0.017 | -0.001 | 0.172 | -0.014 | 0.003 | 0.169 | -0.006 | -0.011 | 0.567 | -0.014 | -0.007 | 0.353 |
|  | **L M1-PM** | 0.028 | 0.060 | 0.104 | 0.043 | 0.069 | 0.142 | 0.104 | 0.106 | 0.928 | 0.099 | 0.051 | 0.005 | 0.041 | 0.015 | 0.016 |
| **2** | **L GP-M1** | -0.035 | -0.004 | 0.131 | -0.026 | 0.001 | 0.056 | -0.018 | 0.004 | 0.096 | -0.024 | -0.014 | 0.330 | -0.003 | -0.007 | 0.625 |
|  | **L GP-PM** | -0.006 | -0.005 | 0.943 | -0.013 | -0.009 | 0.776 | -0.005 | -0.014 | 0.456 | -0.016 | -0.016 | 0.998 | -0.027 | -0.009 | 0.026 |
|  | **L M1-PM** | 0.060 | 0.005 | 0.011 | 0.043 | 0.003 | 0.010 | 0.026 | -0.007 | 0.021 | 0.014 | -0.02 | 0.013 | 0.016 | 0.011 | 0.602 |
|  | **R GP-M1** | -0.103 | 0.007 | 0.003 | 0.140 | -0.015 | <0.001 | 0.118 | -0.023 | <0.001 | -0.048 | -0.02 | 0.072 | -0.130 | -0.029 | <0.001 |
|  | **R GP-PM** | -0.226 | -0.060 | <0.001 | -0.123 | -0.06 | 0.062 | -0.068 | -0.063 | 0.861 | -0.068 | -0.059 | 0.678 | -0.012 | -0.022 | 0.401 |
|  | **R M1-PM** | 0.065 | 0.078 | 0.709 | 0.019 | 0.065 | 0.047 | -0.001 | 0.058 | 0.003 | 0.008 | 0.044 | 0.030 | -0.028 | 0.011 | 0.003 |
| **3** | **L GP-M1** | 0.021 | -0.014 | 0.084 | 0.010 | -0.009 | 0.195 | -0.008 | -0.004 | 0.784 | -0.021 | -0.006 | 0.184 | -0.018 | -0.004 | 0.113 |
|  | **L GP-PM** | 0.020 | -0.017 | 0.028 | 0.026 | -0.015 | 0.003 | 0.018 | -0.014 | 0.033 | -0.005 | -0.009 | 0.704 | 0.001 | -0.008 | 0.323 |
|  | **L M1-PM** | -0.033 | -0.003 | 0.172 | -0.015 | 0.006 | 0.141 | -0.001 | 0.015 | 0.234 | 0.018 | 0.007 | 0.490 | 0.011 | -0.002 | 0.175 |
|  | **R GP-M1** | -0.011 | 0.007 | 0.322 | -0.006 | 0.001 | 0.545 | -0.007 | -0.007 | 0.955 | -0.016 | -0.006 | 0.327 | -0.011 | -0.010 | 0.884 |
|  | **R GP-PM** | 0.014 | 0.001 | 0.481 | 0.020 | 0.002 | 0.158 | 0.022 | 0.002 | 0.065 | -0.003 | 0.002 | 0.596 | -0.003 | 0.001 | 0.629 |
|  | **R M1-PM** | -0.014 | -0.009 | 0.796 | 0.001 | -0.001 | 0.884 | 0.015 | 0.02 | 0.677 | 0.023 | 0.039 | 0.390 | 0.001 | -0.011 | 0.214 |
| **4** | **L GP-M1** | 0.038 | -0.009 | 0.027 | 0.019 | -0.009 | 0.113 | 0.006 | -0.008 | 0.413 | 0.005 | -0.007 | 0.312 | 0.003 | -0.007 | 0.266 |
|  | **L GP-PM** | -0.014 | -0.004 | 0.624 | -0.015 | -0.012 | 0.866 | -0.018 | -0.010 | 0.685 | -0.008 | -0.005 | 0.767 | 0.008 | -0.003 | 0.168 |
|  | **L M1-PM** | -0.042 | 0.040 | 0.004 | -0.026 | 0.015 | 0.031 | -0.008 | 0.005 | 0.464 | -0.013 | 0.037 | 0.016 | 0.001 | 0.009 | 0.427 |

**Supplementary Table S6. Comparison of TRGC scores between in-clinic and at-home recordings |** Mean cortical-pallidal and cortical-cortical TRGC values are shown across delta (δ), theta (Θ), alpha (ɑ), beta (β), and low gamma (γlow) canonical frequency bands for **a**, turning and **b,** straight-walking epochs. Differences between in-clinic and at-home recordings were assessed using permutation testing with Benjamini-Hochberg correction for multiple comparisons. Uncorrected p-values are shown. TRGC, time-reversed Granger causality; L, left; R, right; GP, globus pallidus; M1, primary motor cortex; PM, premotor cortex.

| **Value** | **Region(s)** | **Frequency Band** | **Subject** | | | | | |
| --- | --- | --- | --- | --- | --- | --- | --- | --- |
|  |  |  | **1** | **2** | | **3** | | **4** |
|  |  |  | **L** | **L** | **R** | **L** | **R** | **L** |
| **Power** | **GP** | **δ** | 1 | - | - | - | - | 0.68 |
|  |  | **Θ** | -1 | - | - | - | - | -1 |
|  |  | **ɑ** | 0.51 | - | - | - | - | 0.76 |
|  |  | **β** | -0.38 | - | - | - | - | 0.65 |
|  |  | **γ_low_** | 0.24 | - | - | - | - | 0.66 |
|  | **M1** | **δ** | 0.83 | - | - | - | - | 0.51 |
|  |  | **Θ** | -0.35 | - | - | - | - | -0.21 |
|  |  | **ɑ** | 0.26 | - | - | - | - | 0.56 |
|  |  | **β** | -0.02 | - | - | - | - | 0.04 |
|  |  | **γ_low_** | 0.38 | - | - | - | - | 1 |
|  | **PM** | **δ** | -0.12 | - | - | - | - | 0.51 |
|  |  | **Θ** | 0.31 | - | - | - | - | -0.21 |
|  |  | **ɑ** | 0.36 | - | - | - | - | 0.27 |
|  |  | **β** | -0.85 | - | - | - | - | 0.61 |
|  |  | **γ_low_** | 0.17 | - | - | - | - | 0.33 |
| **Coherence** | **GP-M1** | **δ** | 0.20 | - | - | - | - | 0.20 |
|  |  | **Θ** | 0.01 | - | - | - | - | 0.36 |
|  |  | **ɑ** | -0.01 | - | - | - | - | -0.22 |
|  |  | **β** | 0.07 | - | - | - | - | 0.35 |
|  |  | **γ_low_** | -0.15 | - | - | - | - | 0.07 |
|  | **GP-PM** | **δ** | -0.12 | - | - | - | - | 0.20 |
|  |  | **Θ** | 0.00 | - | - | - | - | 0.09 |
|  |  | **ɑ** | 0.16 | - | - | - | - | 0.42 |
|  |  | **β** | 0.16 | - | - | - | - | 0.27 |
|  |  | **γ_low_** | -0.06 | - | - | - | - | 0.32 |
|  | **M1-PM** | **δ** | -0.03 | - | - | - | - | 0.64 |
|  |  | **Θ** | 0.25 | - | - | - | - | 0.05 |
|  |  | **ɑ** | -0.11 | - | - | - | - | 0.21 |
|  |  | **β** | 0.12 | - | - | - | - | 0.21 |
|  |  | **γ_low_** | -0.17 | - | - | - | - | 0.34 |
| **AUC** | | | 0.61 | 0.54 | 0.56 | 0.52 | 0.60 | 0.68 |
| **p** | | | 0.001 | 0.231 | 0.121 | 0.399 | 0.082 | 0.005 |

**Supplementary** **Table S7. Summary of logistic regression models for classifying turn direction |** Logistic regression models were trained to classify left vs right turns using average power and coherence within canonical frequency bands from GP, M1, and PM signals, separately for each hemisphere. Model coefficients are shown for hemispheres achieving above-chance performance, rescaled from -1 to 1. Mean AUC is reported for each model, with 10-fold cross validation. Empirical one-sided p-values were calculated by randomly shuffling true labels 1,000 times and determining the AUC for each permutation. L, left; R, right; GP, globus pallidus; M1, primary motor cortex; PM, premotor cortex.
